# Evaluation of clinical characteristics of Legionella pneumophila pneumonia diagnosed by metagenomic Next-Generation Sequencing:A Retrospective Study

**DOI:** 10.1101/2024.10.31.24316546

**Authors:** Zhixiong He, Ruixiang Chu, Junyi Ke, Qinglan Li, Qi Wu, Jiao Sheng, Ping Liu, Minchao Duan

**Author notes:** Correspondence: Ping Liu, Department of Respiratory and Critical Care Medicine, The Affiliated Changsha Hospital of Xiangya School of Medicine (The First Hospital of Changsha), Central South University, 311 Yingpan Road, Kaifu District, Changsha City, China. 410000. Correspondence: Minchao Duan, Wuming Hospital of Guangxi Medical University, Wuming District 530100, Nanning, Guangxi, China.

## Abstract

**Purpose:** This study aimed to summarize the clinical characteristics and outcomes of Legionella pneumophila (L. pneumophila) pneumonia to assist clinicians in accurately and early identifying this disease. The findings provide evidence to guide decision-making for L. pneumophila infection and improve treatment success rates.

**Patients and methods:** A retrospective analysis of clinical data was conducted on 19 patients diagnosed with L. pneumophila infection at our hospital using Metagenomic next-generation sequencing (mNGS).

**Results:** The study included 16 male and 3 female participants, with an average age of 59.5±16.2 years. The majority of participants were immunocompromised (13/19). The most common symptoms included fever (n=11) with a median peak value of 39.5°C, cough (n = 15), expectoration (n = 14), dyspnea (n = 10), poor appetite (n = 9), fatigue (n = 8), and headache (n = 3). Elevated levels of C-reactive protein (CRP) and erythrocyte sedimentation rate (ESR) were observed in all patients, while procalcitonin (PCT) and lactate dehydrogenase (LDH) levels were significantly higher in the severe group compared to the non-severe group. Lymphocyte count levels were also significantly lower in the severe group (P<0.05). Common CT manifestations included flake high-density shadows (n = 17), pleural effusion (n = 11), consolidation (n = 4), ground-glass exudation (n = 4), and bronchial inflation signs (n = 3). The most prevalent underlying diseases were hypertension (n = 9), diabetes (n = 8), AIDS (n = 6), COPD (n = 5), and malignancies (n = 3). Common complications included acute liver injury (n=7), acute respiratory failure (n=6), electrolyte disorders (n=5), sepsis (n=3), MODS (n=2), and ARDS (n=1). The majority of patients received treatment with quinolones (16/19), with six patients requiring mechanical ventilation therapy. Tragically, 2 patients in the severe pneumonia group succumbed to sepsis and multiple organ failure.

**Conclusion:** L. pneumophila pneumonia is an acute respiratory infection. If not promptly diagnosed, patients may develop severe pneumonia, multiorgan failure, or face fatal outcomes. mNGS can be used for rapid and accurate diagnosis of L. pneumophila infections, thereby improving treatment outcomes. Early initiation of quinolones or combination therapy with other medications has demonstrated a significant therapeutic benefit for patients with L. pneumophila pneumonia.

## Introduction

Legionella pneumophila is identified as the causative agent of pneumonia, presenting as an acute respiratory infection with potentially fatal consequences (1,2). The transmission primarily occurs through the inhalation of aerosols containing Legionella or inadvertently breathing in water contaminated with L. pneumophila (3). Since the initial discovery of Legionnaires disease in 1976, Legionella pneumophila has increasingly been recognized as a common cause of sporadic and epidemic community-acquired pneumonia (CAP) in individuals of all ages, including both healthy and immunosuppressed individuals (4,5,6). However, it is notably more prevalent in individuals with chronic obstructive pulmonary disease, diabetes mellitus, malignant tumors, uremic patients undergoing extended hemodialysis, organ transplant recipients, and those with compromised immune function(7,8).

Early and rapid diagnosis, as well as pathogen-specific treatment, play a critical role in reducing mortality rates. However, conventional detection methods often encounter obstacles such as challenging cultivation, long detection cycles, and low sensitivity and specificity. The early diagnosis and timely treatment of L. pneumophila pneumonia continue to pose significant clinical challenges. Clinicians often lack awareness of L. pneumophila pneumonia, which is frequently underrecognized or misdiagnosed due to its non-specific symptoms and limited testing options. If patients are not properly diagnosed or if there is a delay in diagnosis, they may develop severe pneumonia, multiorgan failure, or even face fatal outcomes. According to relevant studies, the mortality rate of Legionella pneumonia patients admitted to the ICU can be as high as 33% (9). There is an urgent need for clinical research to better understand the epidemiology and clinical characteristics of L. pneumophila pneumonia. This retrospective study analyzed the clinical data of 19 patients diagnosed with L. pneumophila pneumonia using mNGS. The analysis included demographic characteristics, symptoms, underlying diseases, laboratory data, co-infecting pathogens, computer tomography (CT) imaging, treatment, and outcomes. The purpose of this study is to assist healthcare providers in improving the accuracy and early detection of this disease, providing guidance for clinical management and decision-making related to L. pneumophila infection, and ultimately improving treatment outcomes.

## Material and methods

### Participants

This study was performed by collecting data from medical records of confirmed Legionella pneumophila infection patients at the Affiliated Changsha Hospital of Xiangya School of Medicine between October 1, 2021 and April 30, 2024. Informed consent was obtained from all subjects. The Institutional Review Board (IRB) of The First Hospital of Changsha reviewed and approved our research topic (Grant number: 2024 (11) V. 1.0) in accordance with the ‘Ethical Review Methods for Life Sciences and Medical Research Involving Humans’ (National Health Science and Education Development [2023] No. 4). The authors were unable to obtain information that could identify individual participants during or after data collection. L. pneumophila infection was confirmed by mNGS in bronchoalveolar lavage fluid (BALF), blood, sputum or pleural effusion samples. The inclusion criteria were as follows: (1) patients who met the diagnostic criteria for CAP, (2) L. pneumophila fragments were identified through mNGS analysis of the BALF, blood, sputum or pleural effusion samples, and (3) patients with complete clinical information. In this study, immunocompromised status is defined as any of the following: (1) diabetes mellitus patients; (2) Patients with malignant tumors receiving radiotherapy or chemotherapy; (3) Hematological system Malignant tumors; (4) agranulocytosis after cancer chemotherapy; (5) AIDS patients.

### The mNGS procedures

Metagenomic next-generation sequencing (mNGS) was conducted to detect pathogens in patients with pulmonary infections following established protocols (BGI, Changsha, China). Samples of bronchoalveolar lavage fluid (BALF), sputum, pleural effusion, and blood were collected using standard procedures. BALF or blood samples (3-5 mL) were stored at −20 °C in sterile containers and then sent to BGI (Changsha, China) for analysis. The mNGS process for BALF or blood samples involved nucleic acid extraction, library construction, sequencing, and bioinformatics analysis. Specific steps were not detailed as they were performed by BGI.

### Data collection

Patients’ clinical data included age, sex, underlying diseases, smoking history, laboratory data results, co-infection of principal pathogens, Chest CT images characteristics, treatment protocol, outcomes, and length of hospital stay. Laboratory data were collected before treatment. The data were as follows: white blood cell (WBC) count, neutrophil ratio, lymphocyte count, platelet count, hemoglobin, C-reactive protein (CRP), procalcitonin (PCT), aspartate aminotransferase (AST), alanine transaminase (ALT), erythrocyte sedimentation rate (ESR), lactate dehydrogenase (LDH), creatine kinase (CK), CK-myocardial band (CK-MB) and D-dimer.

### Statistical analysis

Data analysis and mapping were performed by using GraphPad Prism 10. Categorical data were presented as counts and percentages, and measurement data are expressed as mean ± standard deviation (SD) or median with interquartile range (IQR). Comparative analysis was conducted by t-test, and categorical variables were analyzed using Fisher’s exact test. Statistical significance was indicated by p<0.05.

## Results

### Clinical manifestations of patients

Among the 19 patients, 16 were male (84.2%) and 3 were female (15.8%), with an average age of 59.5±16.2 years. The patients were categorized into a severe pneumonia group (n=8) and a non-severe group (n=11), with 6 out of 8 patients (75%) in the severe pneumonia group requiring admission to the ICU. A significant age difference was observed between the severe pneumonia and non-severe groups (p = 0.0108). The median length of hospital stay was 14 days (IQR: 10-23.5). There were no significant differences in sex distribution and length of hospital stay between the severe and non-severe groups. More than half of the patients presented with fever (57.9%, 11/19), with a median peak value of 39.5°C (range 37.8–40.5°C), as well as symptoms such as cough (n = 15, 78.9%), expectoration (n = 14, 73.7%), dyspnea (n = 10, 52.6%), poor appetite (n = 9, 47.4%), fatigue (n = 8, 42.1%), chest tightness (n = 4, 21.1%), headache (n = 3, 15.8%), bloody sputum (n = 1, 5.3%), and myalgia (n = 1, 5.3%). Fatigue was the only clinical symptom that showed a significant difference between the severe and non-severe groups (p = 0.0237). The clinical manifestations of the enrolled patients are detailed in Table 1.

**Table 1.**
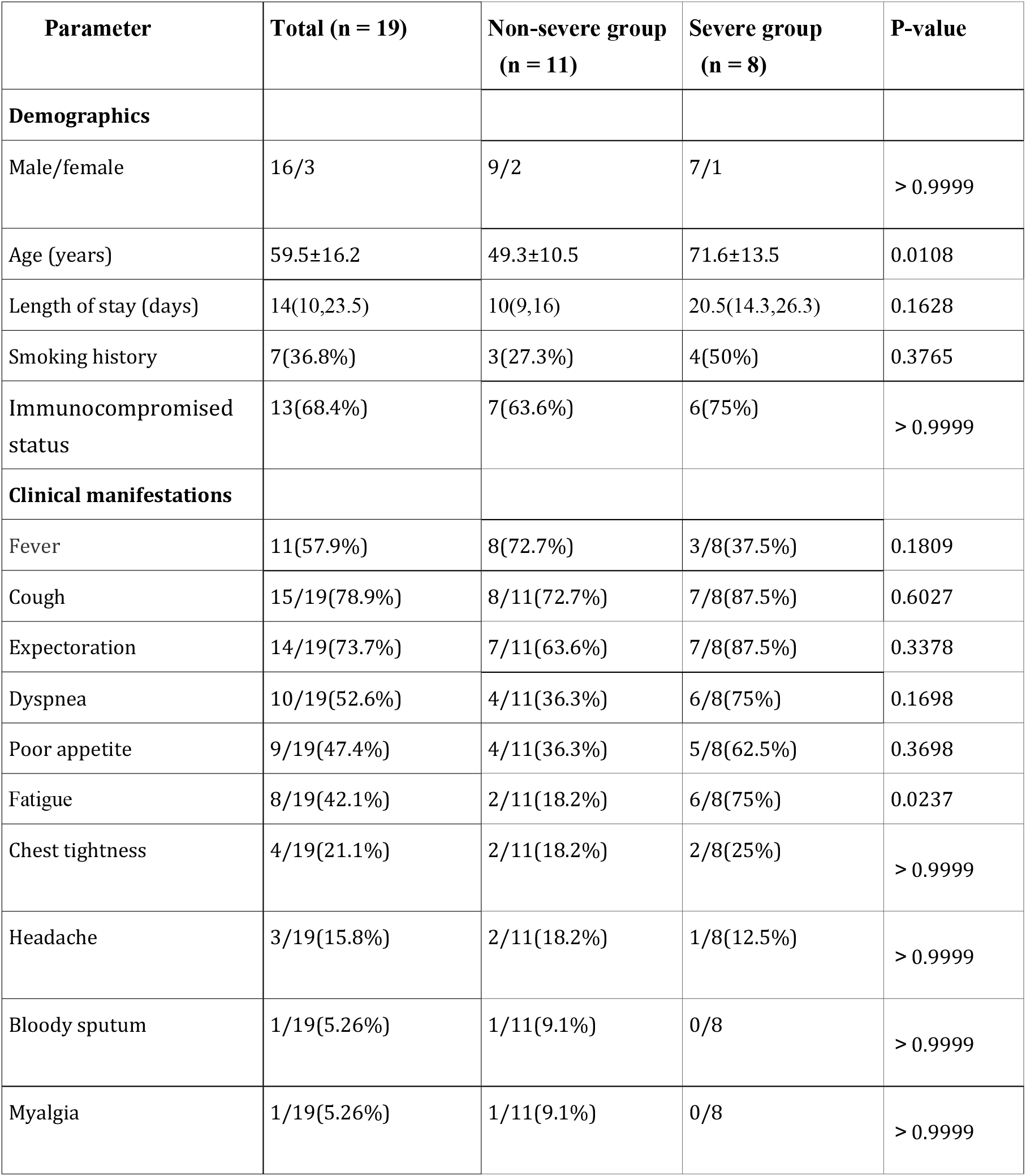
Clinical manifestations of patients with L. pneumophila pneumonia.

### Smoking history, underlying diseases and immune status

Of the 19 patients, nearly half (n=9, 47.4%) were diagnosed in the spring season. Seven patients had a history of smoking (n = 7; 36.8%). The most common underlying diseases were hypertension (n = 9, 47.4%), followed by diabetes mellitus (n = 8, 42.1%). Other common underlying diseases included chronic liver disease (n = 6, 31.6%, encompassing fatty liver and chronic hepatitis B), coronary heart disease (n = 6, 31.6%), AIDS (n = 6, 31.6%), chronic obstructive pulmonary disease (n = 5, 26.3%), chronic renal failure (n = 3, 15.8%), and malignancies (n = 3, 15.8%), such as lung cancer, bladder cancer, and multiple myeloma(Figure 1).

**Figure 1.**
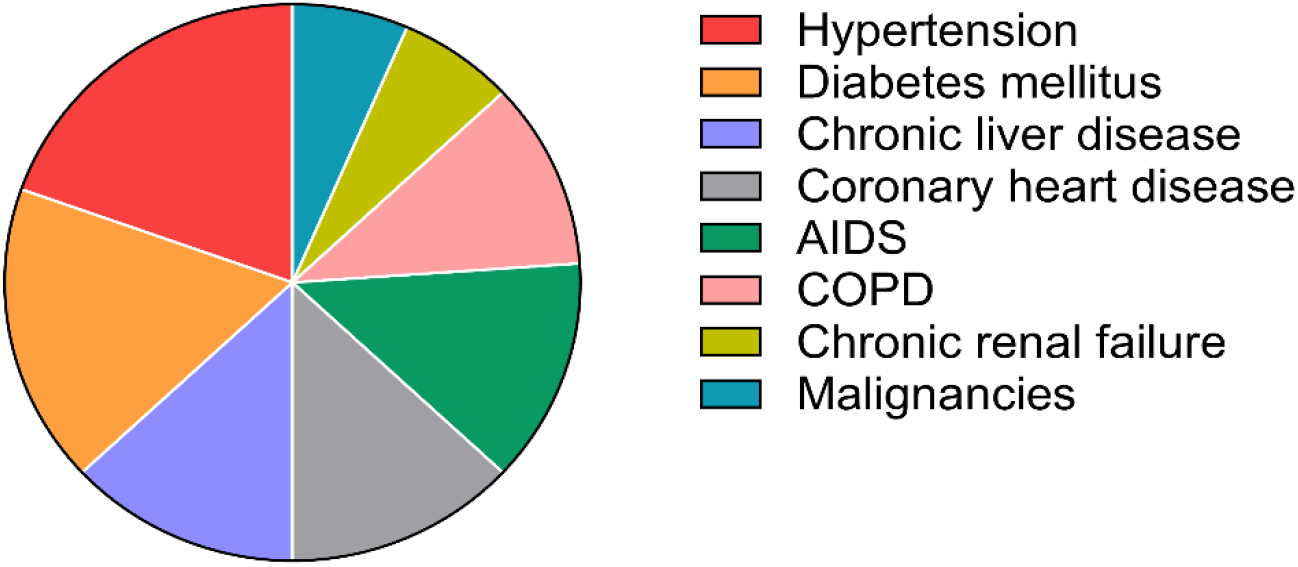
The most common underlying diseases

Immunocompromised patients accounted for the majority (13/19, 68.4%), however, there was no significant difference between the severe group and non-severe group. Among the 13 patients with immunodeficiency, conditions included diabetes mellitus (n=8), AIDS (n=5), lung cancer under radiotherapy and chemotherapy (n=1), multiple myeloma under chemotherapy (n=1), chemoradiotherapy after bladder cancer surgery (n=1), and agranulocytosis after cancer chemotherapy (n=1). Some patients presented with one or more of the aforementioned conditions.

### Laboratory findings and complications

Twelve patients presented with elevated white blood cell (WBC) counts, while only 2 cases had normal WBC counts (both in the non-severe pneumonia group), and five cases had decreased WBC counts (3 cases in the severe pneumonia group and 2 cases in the non-severe pneumonia group) upon admission. Procalcitonin (PCT) and Neutrophil ratio were elevated in all patients in the severe group. C-reactive protein (CRP) and erythrocyte sedimentation rate (ESR) levels were elevated in all patients. PCT levels were found to be >0.05 ng/mL in 15 patients, with four showing an elevation of >2 ng/mL. Additionally, 9 patients had elevated D-dimer levels (mean, 2.05 ± 1.18), and eight patients had elevated LDH levels. Only one patient had an elevated CK-MB level. Both PCT and LDH levels were significantly higher in the severe group compared to the non-severe group (P<0.05), while lymphocyte count levels were significantly lower in the severe group than in the non-severe group (P<0.05), as illustrated in Figure 2-4. The laboratory findings of the two groups are summarized in Table 2. Common complications included acute liver injury (n=7, 36.8%), type I respiratory failure (n=6, 31.6%), electrolyte disorders (n=5, 26.3%), hypoalbuminemia (n=5, 26.3%), sepsis (n=3, 15.8%), acute renal impairment (n=2, 10.5%), MODS (n=2, 10.6%) and ARDS (n=1, 5.3%). (Figure 5).

**Table 2.** Laboratory data of patients with L. pneumophila pneumonia.

| Parameter | Total<br>(n = 19) | Non-severe group<br>(n = 11) | Severe group<br>(n = 8) | P value |
| --- | --- | --- | --- | --- |
| WBC (4.0–10.0×10<sup>9</sup>/L) | 10.67(4.39,13.02) | 10.6(5.91,11.95) | 11.52(3.61,15.5) | 0.7757 |
| NE (50-70%) | 81.3±11.8 | 76.7±11.9 | 87.0±9.3 | 0.0916 |
| LY (1.1-3.2×10<sup>9</sup>/L) | 0.85(0.32, 1.23) | 1.0(0.8, 1.43) | 0.32(0.19, 0.54) | 0.031 |
| PLT (100-300×10<sup>9</sup>/L) | 185.8±91.2 | 207.3±106.4 | 143.5±66.6 | 0.3599 |
| D-Dimer (0-0.5ug/mL) | 1.56(1.17, 2.24) | 1.56 (1.32, 1.61) | 1.6 (1.09, 3.85) | 0.3355 |
| CRP (0-6 mg/L) | 71.37 (35.29, 181.5) | 71.37 (35.3, 175.35) | 94.28 (33.56,192.5) | 0.4321 |
| ESR (0-15mm/h) | 94 (64, 107) | 104(94, 102) | 68 (55, 80) | 0.0683 |
| PCT (0.00-0.05 ng/mL) | 0.49(0.29, 1.78) | 0.49(0.09, 1.35) | 1.35(0.39, 25.77) | 0.0318 |
| LDH (135-225U/L) | 201 (177.1, 292.4) | 184 (172.1, 208) | 269.4(216, 430) | 0.0484 |
| CK (50-310U/L) | 68(33.25, 177.6) | 104 (37.15, 171) | 43.15(32.38, 197.4) | 0.2710 |
| CK-MB (0-25 IU/L) | 9.7 (7.7, 15.1) | 8.2 (7.4, 14.3) | 10.85 (9.6, 17.4) | 0.2235 |
| Albumin(40-55g/L) | 32 (30.35, 35.3) | 33.9 (31.4, 36.05) | 31(28.68, 32.65) | 0.2555 |
| ALT (0-41U/L) | 27.5 (15.1, 36.80) | 27.8 (15.1, 36.75) | 23 (15.7, 50.3) | 0.9824 |
| AST (0-40U/L) | 28.2 (21.9, 58.3) | 28.2 (22.4, 51.2) | 31.5 (22.4, 92.5) | 0.1627 |

**Figure 2-4.**
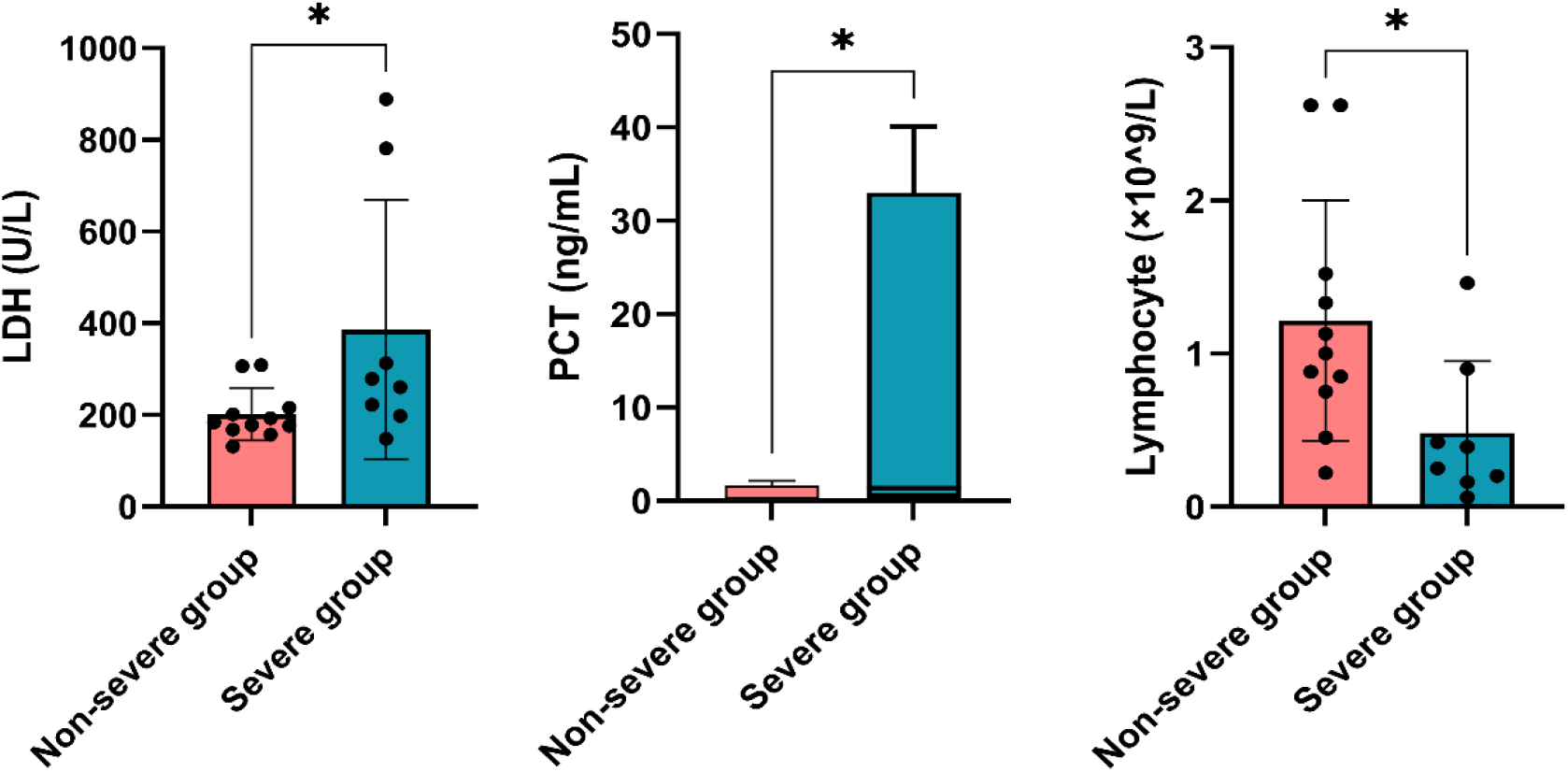
Both PCT and LDH levels were significantly higher in the severe group compared to the non-severe group, while lymphocyte count levels were significantly lower in the severe group than in the non-severe group (P<0.05)

**Figure 5.**
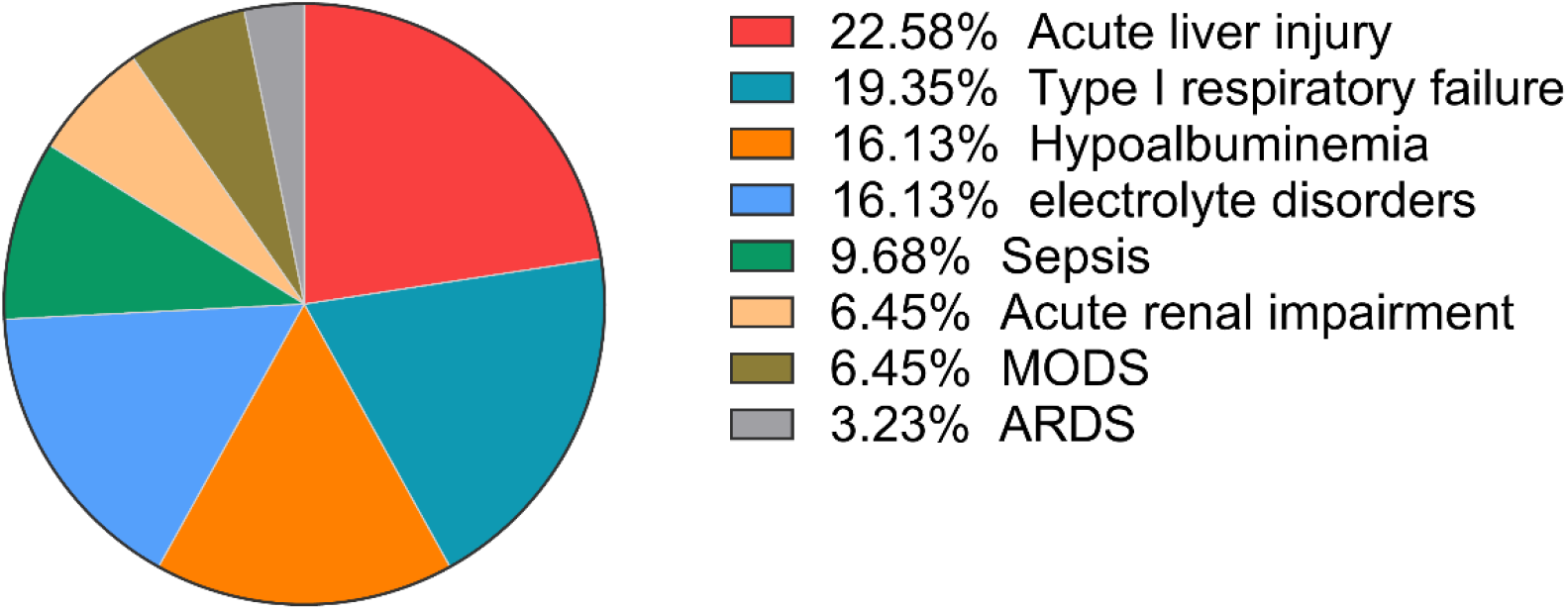
The most common complications

### Chest CT images

All patients in this study underwent chest CT scans, with the most common manifestations being flake high-density shadows (n = 17, 89.5%), pleural effusion (n = 11, 57.9%), small pulmonary nodules (n=5, 26.3%), consolidation (n = 4, 21.1%), ground-glass exudation (n=4, 21.1%) and bronchial inflation signs (n = 3, 15.8%). The distribution of the lesions was as follows: right lung (2/19), left lung (1/19) and both lungs (16/19). Eleven patients had pleural effusion (including six with unilateral pleural effusion and five with bilateral pleural effusion).

### Co-detected pathogens in L. pneumophila pneumonia

In this study, clinical samples identified as L. pneumophila infection using mNGS included 11 samples of BALF, 4 samples of sputum, 3 samples of peripheral blood and 1 sample of pleural effusion. Among these samples, L. pneumophila was identified as the sole pathogen in 5 sample, 73.68% (14/19) patients had mixed infection, including 9 patients with bacterial co-infection, 10 patients with viral infection and 4 patients with fungal infection. 1 cases of co-infection with bacteria and fungi, and 3 cases of co-infection with fungi and viruses. The comprehensive detection rates of bacteria, fungi and viruses were 47.37% (9/19), 21.05% (4/19) and 52.63% (10/19) respectively. The most frequently co-detected pathogen with L. pneumophila was Epstein-Barr virus (7/19), followed by the Cytomegalovirus (5/19). The most common bacterial and fungi co-infection are Staphylococcus aureus and Candida. (Figure 6. a, b).

**Figure 6.a.**
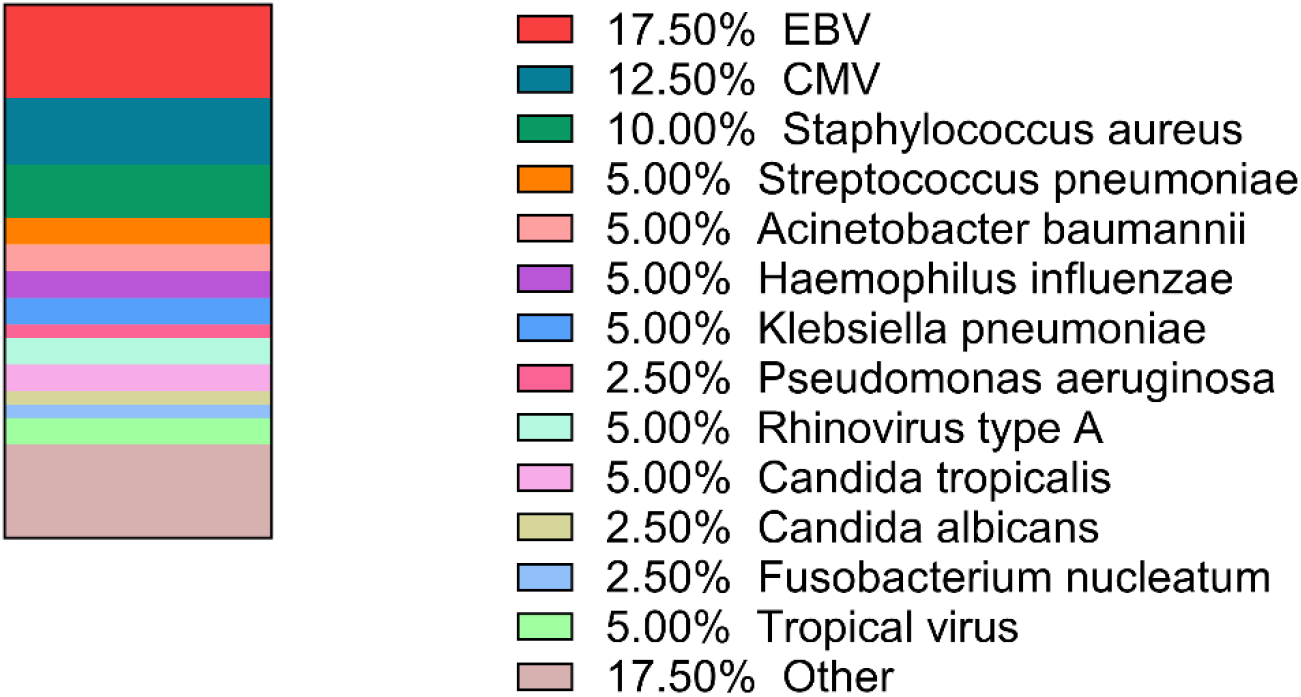
Co-detected pathogens in L. pneumophila pneumonia

**Figure 6.b.**
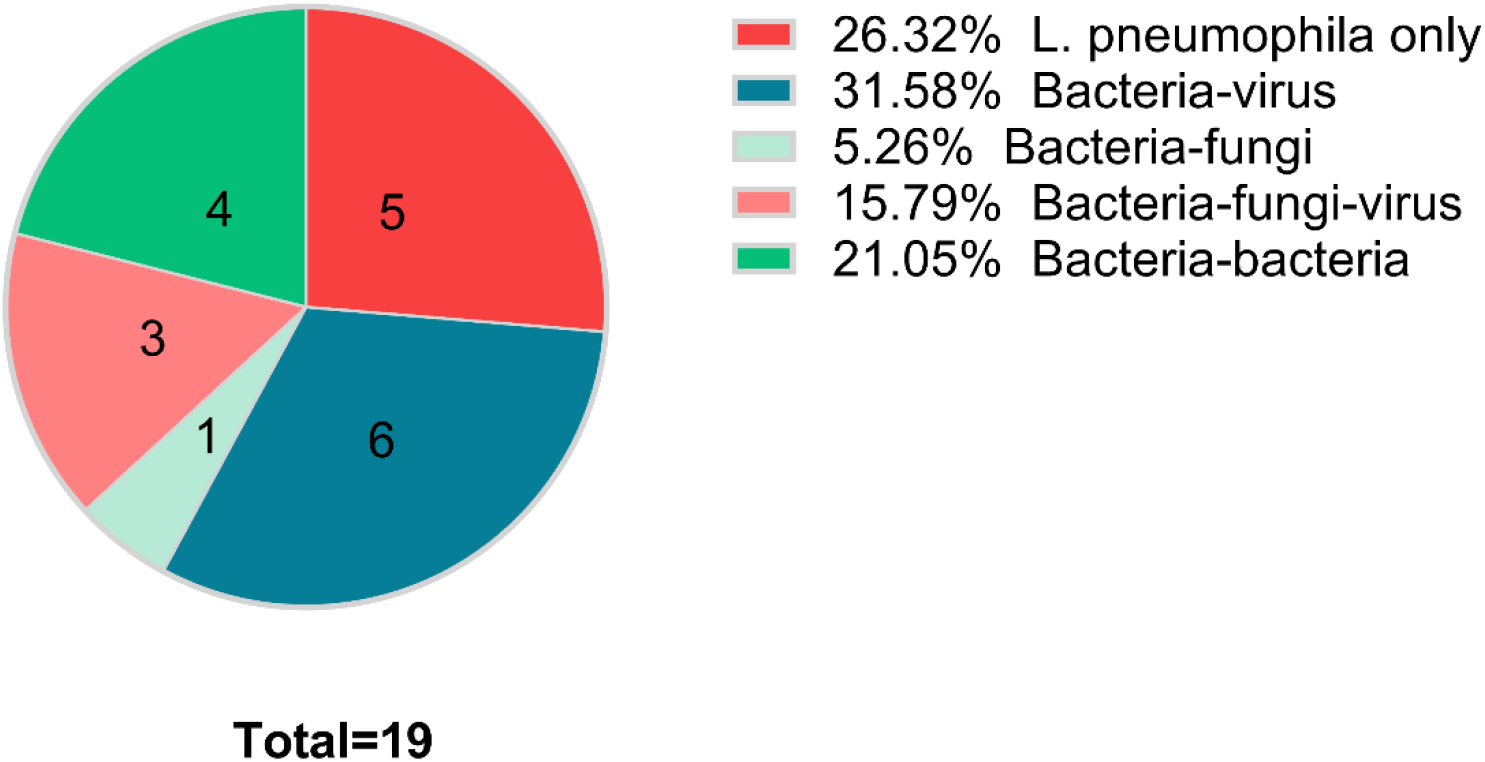
Co-detected pathogens in L. pneumophila pneumonia

### Treatment and outcome

Most patients were treated with quinolones (16/19, 84.2%). Among these patients, 6 of them altered their antibacterial drug regimen based on the mNGS results. Out of the 19 patients, 4 were treated with quinolones alone, 2 with azithromycin and quinolones in combination, 1 with doxycycline and quinolones in combination, 3 received a combination of quinolones and meropenem, 4 patient was treated with a combination of piperacillin/tazobactam and quinolones, 2 were prescribed a combination of quinolones and cefoperazone/sulbactam. Severe group patients were more likely to receive combination therapy compared to non-severe group patients (8/8 vs. 6/11, P = 0.0445). Among the 19 patients, 2 patients were treated with invasive ventilation treatment and 4 patients were treated with non-invasive ventilation treatment in the severe group. Severe group patients were more likely to receive invasive or noninvasive ventilation therapy than non-severe patients (6/8 vs. 0/11, P = 0.001). All patients in the non-severe pneumonia group showed improvement and none of them died during their hospital stay. On the other hand, within the severe pneumonia group, 2 patients passed away due to severe infection and multiple organ failure, while 2 patients were discharged after their condition worsened, with no available follow-up information. The non-severe pneumonia group demonstrated a better prognosis compared to the severe group (11 out of 11 vs. 4 out of 8, P = 0.0181). The in-hospital mortality rate of patients was 10.53% (2 out of 19).

## Discussion

L. pneumophila is a gram-negative, obligate intracellular bacterium that can cause L. pneumophila pneumonia. This disease is more prevalent in America and European countries, with fewer reported cases in China. To our knowledge, this is the largest single-center retrospective observational analysis using mNGS to diagnose patients with L. pneumophila pneumonia in China.

Infection with L. pneumophila typically occurs through inhaling aerosols or water contaminated with the bacteria (3). Previous studies have indicated a correlation between L. pneumophila pneumonia and travel history, with one study in the United States reporting that 20% of legionellosis cases were travel-associated(10,12). However, our study found that none of the patients had a clear travel history. Adults, particularly those with compromised immune systems, are at a higher risk of contracting the disease (12), which is consistent with our study. L. pneumophila is frequently misdiagnosed as a causative agent of severe community-acquired pneumonia (SCAP), and its prevalence is on the rise (13). Some studies have indicated that L. pneumophila pneumonia can occur at any time of the year, but peaks in incidence during the summer and autumn (14,15). However, in our study, nearly half of the cases were diagnosed in the spring season.

This disease is often overlooked due to the limited understanding of the pathogen and the constraints of conventional diagnostic techniques such as culture, urine antigen test, PCR, and serological testing(16,17,18). Additionally, the atypical symptoms of this infection further complicate diagnosis. Once the disease is not diagnosed and treated in time, it may develop into severe pneumonia, sepsis, multiple organ failure, or even death. Fortunately, with the advancements in mNGS, the identification of rare pathogens has become more feasible, leading to better guidance for treatment in clinical settings (19-22).

In this study, the findings revealed that L. pneumophila pneumonia predominantly affects middle-aged and elderly individuals, with a higher susceptibility observed in male patients, and we also found that older patients are more likely to develop severe pneumonia(p<0.05).

Patients with L. pneumophila pneumonia have different disease severity, and their clinical manifestations vary greatly. Common clinical manifestations observed in patients with this condition included high fever, cough, expectoration, fatigue, poor appetite, and dyspnea. Severe cases often experienced respiratory failure, septic shock, and organ dysfunction. The clinical presentations of the patients in this study closely aligned with those reported in previous research (7).

In our study, twelve patients presented with elevated white blood cell (WBC) counts, only 2 cases presented with normal WBC counts, and five cases presented with decreased WBC counts. CRP and ESR levels were elevated in all patients, PCT and Neutrophil ratio were elevated in all patients of the severe group. Only one patient presented with elevated CK-MB level. Acute liver injury, acute respiratory failure, electrolyte disorders and hypoalbuminemia is common, which is consistent with previous study (23). The PCT and LDH levels were significantly higher in the severe group than in the non-severe group(P<0.05), and lymphocyte count levels were significantly lower in the severe group than in the non-severe group(P<0.05), perhaps we can use lymphocyte count indicators to assess disease severity and prognosis. Moreover, we know that lymphocytes are related to the body’s immune function, which further suggests that the occurrence and severity of this disease are related to the patient’s immune status.

Our study showed that chest CT plays an important role in diagnosing L. pneumophila pneumonia. The major imaging manifestations include flake high-density shadows, pleural effusion, some may be accompanied by consolidation, ground-glass exudation, small nodules and air bronchus signs. Our study identified small pulmonary nodule changes on CT scans in some patients, a phenomenon that has been rarely reported in previous studies. We hypothesize that these changes may be linked to pulmonary inflammation in patients, although further research is needed to explore the specific reasons behind this relationship. However, necrosis, cavity, and tree-in-bud signs were not observed, and this could help in distinguishing L. pneumophila infection from other infections.

The mNGS results confirmed the presence of mixed infections in our study, with 73.7% of the patients exhibiting this phenomenon. The most common co-infections were found to be associated with Epstein-Barr virus, followed by cytomegalovirus. The most common bacterial and fungi co-infection are staphylococcus aureus and candida. It was observed that co-infections are more likely to occur in the severe pneumonia group, indicating that co-infections in patients with L. pneumophila pneumonia could lead to more severe infections and a poorer prognosis (23), highlighting the need for further research in this field.

A respiratory quinolone and/or a macrolide are the recommended treatments for L. pneumophila pneumonia, with Levofloxacin or azithromycin being the first-line drugs (25,26). In this study, most patients were treated with quinolones (84.2%) alone or in combination for initial antimicrobial treatment, with most experiencing positive therapeutic outcomes. Some patients initially had poor responses, prompting treatment adjustments following clear pathogen identification via mNGS. Upon switching to quinolone or a quinolone combination, most of the patients showed improvement. Our findings suggest that Quinolones were effective in treating L. pneumophila pneumonia, highlighting the efficacy of quinolones. Further clinical studies are warranted to confirm the effectiveness of these drugs against L. pneumophila infections.

In a previous study, the mortality rate among Legionella pneumonia cases admitted to the ICU was approximately 33%. In our study, the mortality rate among L. pneumophila pneumonia cases admitted to the ICU was slightly lower at 25%. This difference may be attributed to the exclusion of two patients from the mortality calculation, who were discharged after their condition deteriorated. An additional study has shown that initiating fluoroquinolone therapy promptly can result in a decrease in mortality rates. When the correct antibiotics are administered early, mortality decreases to below 5%. On the contrary, a delay in initiating the appropriate antibiotics is associated with a worse prognosis (27,28).

Although our study provided valuable insights, it was constrained by several limitations. The small sample size of only 19 patients restricted our ability to comprehensively investigate all relevant characteristics of L. pneumophila pneumonia. Additionally, as a single-center retrospective study, there may have been a presence of selection bias among participants. Furthermore, all cases included in the study were diagnosed exclusively through mNGS, without incorporating other diagnostic methods such as serological tests, PCR, and/or culture.

## Conclusion

L. pneumophila is a common causative pathogen in hospitalized patients with CAP, often leading to high mortality rates in cases of severe pneumonia. This study aimed to evaluate the characteristics of L. pneumophila pneumonia, treatment strategies, and outcomes. Clinicians are advised to consider L. pneumophila infection in cases of atypical pneumonia presenting with relevant clinical symptoms and abnormal findings on Chest CT scans. Empirical treatment with quinolones is recommended for patients with severe atypical pneumonia. mNGS can assist in early diagnosis and the prompt initiation of effective treatment.

## Abbreviations

L. pneumophila: Legionella pneumophila
mNGS: metagenomic next-generation sequencing
CAP: community-acquired pneumonia
BALF: bronchoalveolar lavage fluid
NE: neutrophil
LY: lymphocyte
CK: creatine kinase
CRP: C-reactive protein
ESR: erythrocyte sedimentation rate
PCT: procalcitonin
ARDS: acute respiratory distress syndrome
MODS: multiple organ failure syndrome

## Competing Interests

The authors have no relevant financial or non-financial interests to disclose.

## Author Contributions

Zhixiong He, Junyi Ke and Qinglan Li participated in the data sorting, analysis, and drafting of the manuscript. Ping Liu and Minchao Duan participated in the data analysis and revised the manuscript. Ruixiang Chu, Qi Wu and Linyan Xu participated in data collection, sorting and analysis. Jiao Sheng participated in data collection and sorting. All authors approved the submitted version.

## Data Availability

The data analysed during the current study are available in the Harvard Dataverse (https://doi.org/10.7910/DVN/SFXXBM).

### Compliance with ethical standards

This study was performed in line with the principles of the Declaration of Helsinki. The study was approved by the Ethics Committee of The Affiliated Changsha Hospital of XiangyaSchool of Medicine (Grant number: 2024 (11) V. 1.0). The authors were unable to obtain information that could identify individual participants during or after data collection. The Institutional Review Board (IRB) of The First Hospital of Changsha reviewed and approved our research topic in accordance with the ‘Ethical Review Methods for Life Sciences and Medical Research Involving Humans’ (National Health Science and Education Development [2023] No. 4).

## Acknowledgements

We thank the patients and their families for their support of this study. We thank BGI(Changsha) for their detection technical assistance.

## Notes

### Competing Interest Statement

The authors have declared no competing interest.

### Funding Statement

The author(s) received no specific funding for this work.

### Author Declarations

The study was approved by the Ethics Committee of The Affiliated Changsha Hospital of XiangyaSchool of Medicine (Grant number: 2024 (11) V. 1.0).

